# Heparin Does Not Regulate Circulating Human PCSK9

**DOI:** 10.1101/2022.09.12.22279796

**Authors:** Vivian Q. Xia, Chui Mei Ong, Lucas S. Zier, John S. MacGregor, Alan H. B. Wu, John S. Chorba

**Affiliations:** Division of Cardiology, Zuckerberg San Francisco General Hospital; Department of Medicine, University of California San Francisco; Clinical Chemistry Laboratory, Zuckerberg San Francisco General Hospital; Department of Laboratory Medicine, University of California San Francisco

## Abstract

**Background:** PCSK9 chaperones the hepatic low-density lipoprotein receptor (LDLR) for lysosomal degradation, elevating serum LDL cholesterol and increasing the risk of atherosclerotic heart disease. Though the major effect on the hepatic LDLR comes from secreted PCSK9, the details of PCSK9 reuptake into the hepatocyte remain unclear. In both tissue culture and animal models, heparan sulfate proteoglycans (HSPGs) on hepatocytes act as co-receptors to promote PCSK9 reuptake. We hypothesized that if this PCSK9:HSPG interaction is important in humans, disrupting the interaction with unfractionated heparin (UFH) would acutely displace PCSK9 from the liver and increase plasma PCSK9.

**Methods:** We obtained remnant plasma samples from 160 subjects undergoing cardiac catheterization before and after administration of intravenous UFH. PCSK9 levels were determined using a commercial ELISA.

**Results:** Median plasma PCSK9 concentrations were 113 ng/ml prior to UFH and 119 ng/ml afterwards. This difference was not significantly different (*p* = 0.83, 95% CI of difference: −6.23 to 6.31 ng/ml). Tests for equivalence provided 95% confidence that UFH administration would not raise PCSK9 levels by more than 4.7% of the baseline value. No predefined subgroups exhibited an effect of UFH on circulating PCSK9.

**Conclusion:** Administration of UFH does not result in a clinically meaningful effect on circulating PCSK9 in humans. These results suggest that disruption of the PCSK9:HSPG interaction does not affect PCSK9 reuptake into the liver. Further, they cast doubt on the clinical utility of disrupting the PCSK9:HSPG interaction as a therapeutic strategy for PCSK9 inhibition.

**Graphical Abstract:** 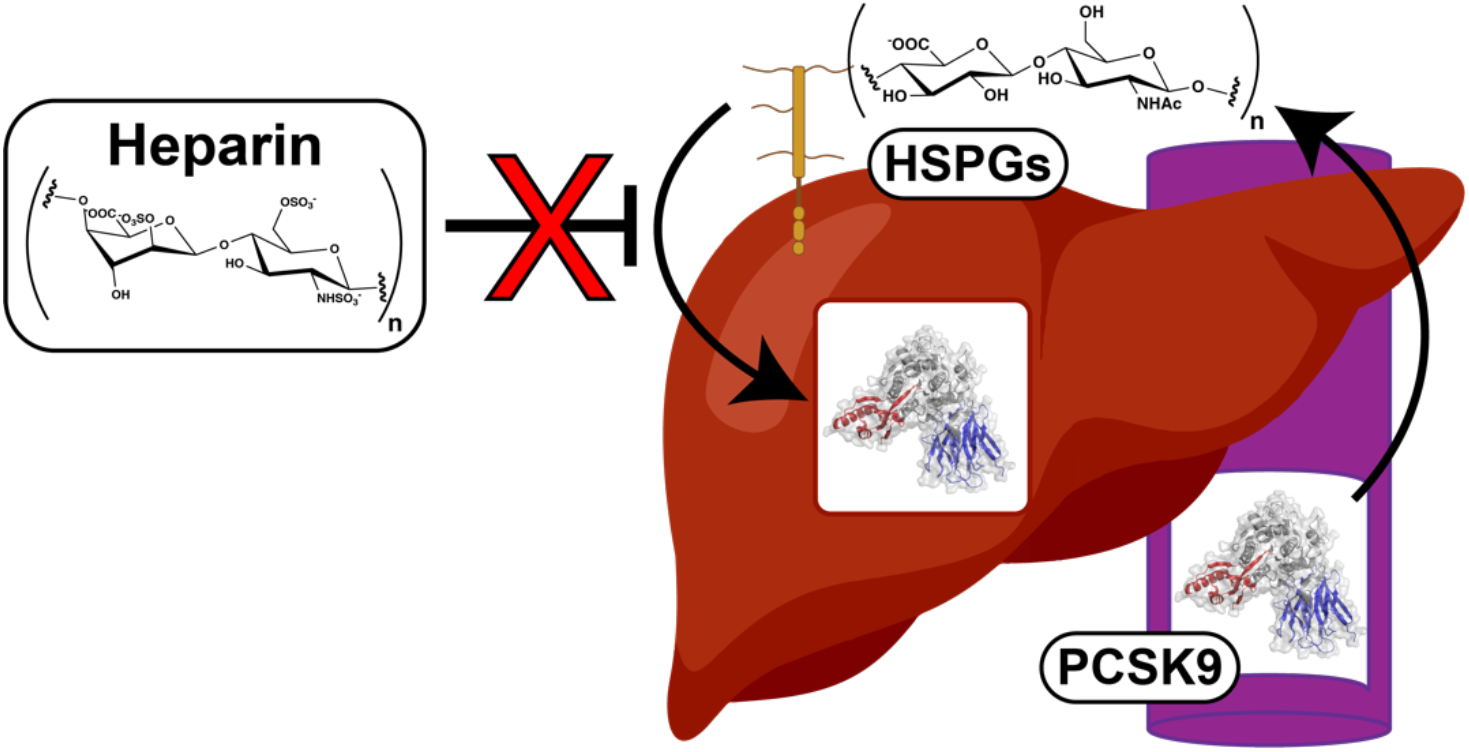

**Clinical Perspective:** *What is new?:*

- Prior tissue culture and animal data suggest that PCSK9 interacts with hepatic heparan sulfate proteoglycans to enter the liver and raise cholesterol levels
- We found no evidence that heparin, a competitive inhibitor of heparan sulfate proteoglycans, acutely affects PCSK9 in humans

*What are the clinical implications?:*

- Heparin is unlikely to be successfully repurposed as a PCSK9 inhibitor

## Main Text

PCSK9 chaperones the hepatic low-density lipoprotein receptor (LDLR) for lysosomal degradation, elevating serum LDL cholesterol and increasing the risk of atherosclerotic heart disease^1^. Though the major effect on the hepatic LDLR comes from secreted PCSK9^2^, the details of PCSK9 reuptake into the hepatocyte remain unclear. In both tissue culture and animal models, heparan sulfate proteoglycans (HSPGs) on hepatocytes bind to an arginine-rich region on the PCSK9 prodomain to promote PCSK9 reuptake^3,4^. However, to date, the PCSK9:HSPG interaction has not been directly evaluated in humans. We hypothesized that if this interaction is important, disrupting it with unfractionated heparin (UFH) would acutely displace PCSK9 from the liver and increase plasma PCSK9. We therefore leveraged remnant blood samples from patients undergoing cardiac catheterization and short-term anticoagulation with intravenous (IV) UFH in the <u>Hep</u>arin <u>Block</u>ade of PCSK<u>9</u> (“HepBlock9”) study (Fig. 1A). To our surprise, we observed no difference in circulating PCSK9 after UFH administration, suggesting that heparin is unlikely to be successfully repurposed for PCSK9 inhibition.

**Figure 1:**
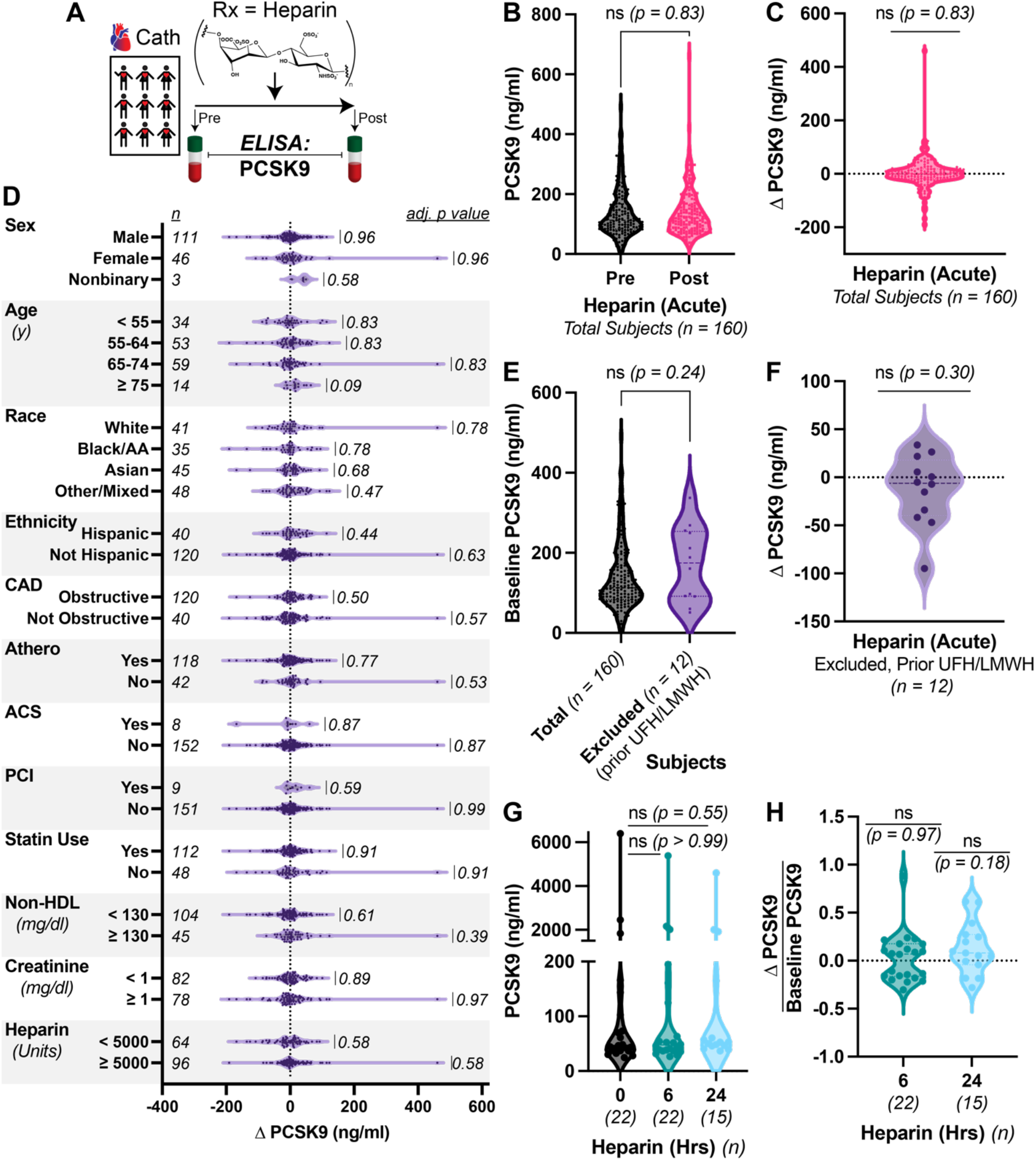
HepBlock9 Study. *A)* Schematic of study. Remnant blood samples before and after heparin administration in subjects undergoing cardiac catheterization were tested for PCSK9 concentrations. *B)* Plasma PCSK9 concentrations in study subjects before (black) and after (pink) IV UFH administration. *C)* Difference in plasma PCSK9 concentrations before and after IV UFH in total subjects. Source data is the same as from *B. D)* Difference in plasma PCSK9 concentrations before and after IV UFH in subjects stratified by sex, age, race, ethnicity, obstructive CAD, atherosclerosis, ACS presentation, percutaneous coronary intervention (PCI), statin use, serum non-HDL cholesterol, serum creatinine, or heparin dose. *E)* Baseline (pre-UFH) plasma PCSK9 concentrations in study subjects (after exclusions, black) and subjects excluded due to prior UFH or low-molecular-weight heparin (LMWH) administration (purple). Mann-Whitney test shown. *F)* Difference in PCSK9 concentrations before and after IV UFH in subjects excluded due to prior UFH or LMWH administration. *G)* Plasma PCSK9 concentrations before (black) or after 6 (teal) or 24 (light blue) hr infusions of IV UFH. Note the discontinuous Y axis. *H)* Fractional difference in plasma PCSK9 concentrations before and after IV UFH infusions. Source data is the same as from *G. All panels)* Unless otherwise noted, Wilcoxon signed-rank tests shown, comparing matched-pairs between groups or to a hypothetical median of 0, with Holm-Sidak corrections for multiple hypothesis testing when appropriate. *n* = number of subjects in each group, ns = non-significant at adjusted *p* > 0.05.

In a protocol approved with a waiver of informed consent by the Institutional Review Board of the University of California, San Francisco (UCSF, CHR #18-26734), we targeted a sample size of 150 subjects for 80% power at α = 0.05 (two-tailed) to detect a 4 ng/ml (7%) change in PCSK9^5^. This was a conservative calculation given the 25% reduction in PCSK9 observed upon PCSK9:HSPG disruption in animal models^3^. We then collected 180 paired pre- and post-UFH remnant plasma samples, excluding 20 subjects for unclear records of UFH administration, a prolonged (> 8 hr) delay in sample collection, recent heparin use, or evidence of impaired liver function. This left us with 160 subjects available for analysis, who represented a predominantly male (69%) but ethnically diverse cohort (74% non-White) with the expected high prevalence of atherosclerosis (74%), statin use (70%), and obstructive coronary disease (40%).

We then measured plasma PCSK9 concentrations using a commercial ELISA (Abcam) compatible with our collection strategy and reviewed the associated medical records. Median PCSK9 concentrations were 113 ng/ml prior to UFH and 119 ng/ml after, with no significant difference between the matched samples (*p* = 0.83, Fig. 1B&C). Equivalence testing provided 95% confidence that the rise in PCSK9 from UFH was no more than 5.35 ng/ml, or 4.7% of the baseline. Because several homeostatic mechanisms regulate circulating PCSK9^5^, we also evaluated the effect of IV UFH on circulating PCSK9 after stratifying by sex, age, race, ethnicity, burden of atherosclerosis, presentation of acute coronary syndrome (ACS), statin use, non-HDL cholesterol, renal function, and UFH dose. However, we failed to identify any subgroup that displayed a significant response (Fig. 1D). We conclude from these data that administration of IV UFH does not raise PCSK9 concentrations acutely.

We focused on bolus administration of UFH because the reported half-life of circulating PCSK9 is only 5 minutes^2^. To assess whether longer UFH treatments are required to disrupt PCSK9 uptake, we compared baseline PCSK9 concentrations in our main cohort to the 12 subjects excluded because of prior treatment with heparin-like products, but found no significant difference (*p* = 0.24, Fig. 1E&F). We also measured PCSK9 concentrations in 22 separate patients anticoagulated with continuous IV UFH. Median PCSK9 concentrations in the paired samples were no different after either a 6 hr (*p* > 0.99, Fig. 1G) or 24 hr infusion (*p* = 0.55, Fig. 1G). Because this cohort displayed a wide distribution of baseline PCSK9 concentrations, we also explored the effects on the fractional change in PCSK9 at both timepoints, but again found no significant difference (*p* = 0.97 for 6 hrs, *p* = 0.18 for 24 hrs, Fig. 1H).

Together, our data support the null hypothesis: that administration of therapeutic UFH does not disrupt PCSK9 reuptake or affect circulating PCSK9 concentrations. Our results highlight the importance of assessing physiologic questions in humans to assess their clinical relevance, given the observations on the PCSK9:HSPG interaction in prior models^3,4^. The strengths of our study include 1) direct measurements in human subjects, without need for an experimental animal model, 2) the clinical relevance to potential drug repurposing, even if the answer is negative, 3) temporal observations to draw causal inferences, 4) the diverse ethnic population of our sample, improving the generalizability of our findings, and 5) the use of subjects as their own controls to minimize confounding from the substantial variability in PCSK9 concentrations among individuals. Our study is limited by 1) a reliance on static measurements, leaving us unable to directly assess PCSK9 kinetics or shifts between body compartments, 2) a sample size powered for our main hypothesis but not necessarily subgroup analyses, and 3) the focus on acute UFH administration, which limits our conclusions regarding long-term HSPG blockade. Given the relative infrequency of PCSK9 variants, we also cannot comment on the role of the PCSK9:HSPG interaction in certain genetic backgrounds, which may yet remain important^4^.

In conclusion, we found no evidence that heparin acutely interferes with PCSK9 uptake, casting doubt on the clinical utility of disrupting the PCSK9:HSPG interaction as a potential strategy to inhibit PCSK9 function.

## Supporting information

STROBE Checklist

## Data Availability

Additional data and materials requests are available from the corresponding author via material transfer agreement, subject to IRB review.

## Acknowledgments

We thank the UCSF cardiology fellows and nursing staff of the ZSFG cardiac catheterization laboratory for preservation of remnant samples.

## Funding

This work was supported by the NIH/NHLBI (K08 HL124068, R03 HL145259, R01 HL146404, and R01 HL159457), a Pfizer ASPIRE Cardiovascular Award, the Harris Fund, the Research Evaluation and Allocation Committee of the UCSF School of Medicine, all to J.S.C.

## Author contributions

Overall study design: J.S.C; Supervision of preservation of remnant samples: L.S.Z, J.S.M.; Storage and processing of remnant samples: C.M.O, A.H.B.W.; Determination of PCSK9 concentrations: V.Q.X, J.S.C; Data analysis: V.Q.X, J.S.C; Manuscript preparation: J.S.C; Critical review and revision of the manuscript: All authors.

## Competing interests

J.S.C. has received consulting fees from Gilde Healthcare and Eko.

## Notes

### Author Declarations

The Institutional Review Board of the University of California, San Francisco (UCSF) gave ethical approval for this work with a waiver of informed consent (CHR #18-26734).

