## Supplemental Methods for "Heparin Does Not Regulate Circulating Human PCSK9"

Extended Methods

*Study Subjects and Samples*

All protocols were approved by the Institutional Review Board of the University of California, San Francisco (UCSF) as a no subject contact study with a waiver of informed consent (CHR #18-26734). Adult subjects presenting to the cardiac catheterization laboratory at the Zuckerberg San Francisco General Hospital (ZSFG) receiving intravenous (IV) UFH were considered. Remnants from blood samples routinely obtained during the clinical procedure were transferred to sterile heparinized tubes by procedural staff and promptly placed on ice. Within 8 hrs, study personnel collected the sample, isolated the plasma by centrifugation (1000 × *g* for 10 min at 4 °C), and re-labeled and stored the deidentified samples at -80 °C for later biochemical analysis. Based on the standard deviation ($\sigma$) of plasma PCSK9 in the general population, we estimated that 150 subjects would provide 80% power ($1 - \beta$) at 95% confidence (two-tailed $\alpha$ = 0.05) to detect a 4 ng/ml change in PCSK9 concentrations (7% of total) from UFH administration. This effect size is a conservative estimate compared to the 25% increase in circulating murine PCSK9 observed after heparinase treatment.

In a separate exploratory cohort, we also obtained samples from inpatient subjects receiving a continuous infusion of IV UFH. Remnants from citrated blood samples, which were used to determine the partial thromboplastin time (PTT) before and 6 hrs after initiation of UFH, were collected by study personnel after room temperature storage in the clinical laboratory for up to 24 hrs. Plasma was isolated, re-labeled, deidentified, and stored at -80 °C as noted above.

Clinical data, including demographics, clinical laboratory values, medication records, medical histories, diagnosis codes, and cardiac catheterization reports, were obtained from the longitudinal electronic medical record and reviewed by study investigators remote from the time of clinical presentation.

*Inclusion and Exclusion Criteria*

In the main cohort, we included all adults ≥ 18 years of age presenting to the cardiac catheterization laboratory who had blood samples obtained immediately before and approximately 5 to 20 minutes after receiving IV UFH for their procedure. Exclusion criteria were limited to 1) active use of heparin or heparinoids, defined as IV UFH within 4 hrs of the procedure, subcutaneous (SC) UFH or prophylactic dosing of SC low-molecular weight heparin (LMWH) within 24 hrs of the procedure, or therapeutic dosing of SC LMWH within 48 hrs of the procedure, or 2) active liver disease, defined by plasma alanine aminotransferase (ALT), aspartate aminotransferase (AST), alkaline phosphatase (AP), or total bilirubin (TB) greater than three times the upper limit of normal.

Pre-defined strata included the following variables: 1) sex, defined as male, female, or nonbinary, with the latter encompassing all subjects with any discordance between legal sex, sex at birth, or gender identity; 2) age, defined as <55, 55-64, 65-74, or ≥75 years; 3) race, self-identified as White, Black or African American, Asian, American Indian or Native Alaskan, Native Hawaiian or other Pacific Islander, Other, Unknown; 4) ethnicity, self-defined as Hispanic or not Hispanic; 5) obstructive coronary artery disease (CAD), defined as the presence of an ≥70% stenosis in any coronary artery upon angiography, a ≥50% stenosis in the left main coronary artery, or evidence of stenosis by physiologic testing such as fractional-flow reserve (FFR) or instant wave-free ratio (iFR); 6) atherosclerosis, defined as the presence of any atherosclerotic plaque on coronary angiography; 7) acute coronary syndrome (ACS), defined as evidence of an acute plaque rupture, thrombus, or culprit lesion on coronary angiography; 8) percutaneous coronary intervention (PCI) performed during cardiac catheterization; 9) active statin use at time of cardiac catheterization; 10) non-HDL cholesterol; 11) creatinine; and 12) dose of IV UFH during cardiac catheterization. All clinical definitions in the record were adjudicated by a board-certified cardiologist.

*Enzyme-Linked Immunosorbent Assays (ELISAs)*

Upon collection of sufficient samples to run an entire plate, blood samples were thawed and analyzed via a commercial PCSK9 ELISA kit (Human PCSK9 SimpleStep ELISA, ab209884, Abcam) compatible with heparinized samples according to the manufacturer's instructions with minor modifications. Samples were diluted 50-fold in sample diluent and assayed in duplicate, with all measurements resulting within the reference range of the manufacturer. End point absorbance at $\lambda$ = 450 nm was recorded on a Spark plate reader (Tecan). Plasma PCSK9 concentrations were interpolated from a 4-parameter logistic regression curve (Prism 9, GraphPad Software) fit to the background-subtracted PCSK9 standard run in each plate. For the exploratory cohort of subjects treated with UFH for 6 hrs, a commercial ELISA compatible with citrated samples was used (RayBiotech, ELH-PCSK9).

*Statistical Analysis*

Interpolated PCSK9 concentrations from pre- and post-heparin samples were tested for normality using the D'Agostino and Pearson test. For non-Gaussian data, pairwise comparisons were performed with nonparametric tests: the Mann-Whitney test was used to compare between two groups with unpaired data, the Wilcoxon matched-pairs signed rank tests was used for paired data, and the Kruskal-Wallis and Dunn's multiple comparisons tests were performed to compare among three or more groups. The Wilcoxon signed rank test was used to compare the change in PCSK9 levels to the hypothetical median of 0. When appropriate, the Holm-Sidak correction was applied to account for multiple strata. For Gaussian data, pairwise comparisons were performed with paired t-tests. Statistical significance was set at adjusted $p$ = 0.05.
